# A Case-Control Study of the Association Between *Karenia Brevis* (Red Tide) and Biliary Atresia

**DOI:** 10.1101/2022.10.24.22279447

**Authors:** Rita Wyrebek, Jamie L Fierstein, Rebecca G. Wells, Joana Machry, Sara Karjoo

**Affiliations:** Saint Louis University, Department of Pediatrics, Division of Neonatology, 1465 S Grand Blvd, St. Louis, MO 63104; Johns Hopkins All Children’s Hospital, Institute for Clinical and Translational Research, Epidemiology and Biostatistics Shared Resource, 501 6^th^ Ave S, St. Petersburg, FL 33701; University of Pennsylvania, Division of Gastroenterology and Hepatology, 905 BRB II/III, 421 Curie Blvd., Philadelphia, PA 19104; Johns Hopkins All Children’s Hospital, Department of Maternal, Fetal and Neonatal Medicine, Division of Neonatology, 501 6^th^ Ave S, St. Petersburg, FL 33701; Johns Hopkins All Children’s Hospital, Division of Gastroenterology, 501 6^th^ Ave S, St. Petersburg, FL 33701

**Keywords:** neonatal cholangiopathy, brevetoxin, environmental toxins, algal bloom, gestational exposure

## Abstract

**Objective:** The study objective was to evaluate the association between maternal *Karenia brevis (K. brevis)* exposure during pregnancy and the prevalence of biliary atresia (BA) in offspring. Study Design This was a hospital-based, case-control study in which cases were infants diagnosed with BA at Johns Hopkins All Children’s Hospital from October 2001 to December 2019. Controls were matched 4:1 by age, randomly selected from healthy infants hospitalized during the study period for common pediatric diagnoses. Infants were excluded if they had congenital anomalies and/or were non-Florida residents. Gestational *K. brevis* exposure levels (cells/liter) were determined from Florida Fish and Wildlife Conservation Commission exposure data at 10- and 50-mile radii from the mother’s zip code of residence. Multivariable conditional logistic regression determined adjusted odds of BA in offspring based on maternal gestational *K. brevis* exposure.

**Results:** Of 38 cases and 152 controls, no significant inter-group differences were observed in race/ethnicity, season of birth or coastal residence. Median gestational exposure at the 10-mile radius was 0 cells/liter in both groups. A greater proportion of cases had no *K. brevis* exposure (63.2%, n = 24) in comparison to controls (37.5%, n = 57; p = .04) at a 10-mile radius. At the 50-mile radius, cases had a peak median exposure at 6 months of gestation compared to controls’ peak at 9 months. After adjustment for sex, seasonality, race/ethnicity, and coastal residence, there was no significant association between BA and maximum *K. brevis* exposure per trimester of pregnancy at the 10-or 50-mile radius.

**Conclusion:** We observed no association between gestational *K. brevis* (cells/liter) exposure at a 10- and 50-mile radius from maternal zip code of residence and BA in offspring.

**Key points:**

- Environmental toxins may cause biliary atresia (BA)
- Red tide is caused by algal blooms (*Karenia brevis)*
- Red tide is debilitating to marine wildlife
- Gestational exposure to *K. brevis* does not cause BA

## INTRODUCTION

Prenatal exposure to environmental toxins ^1–5^ is one of the possible mechanisms underlying the development of biliary atresia (BA), a devastating, progressive neonatal cholangiopathy characterized by extra-hepatic bile duct obstruction ^6^. Liver development begins as early as the second week of embryonic life with bile ducts appearing by the 8^th^ week of life ^7,8^, during which time the fetus is especially vulnerable to toxic insults.

The underlying pathophysiology of BA is unknown, with many proposed etiologies (genetic, infectious, toxin-mediated). Environmental toxins have been previously implicated in BA development in animals ^1–4^. Recently, a plant isoflavonoid named biliatresone was found to cause a BA-like disease in livestock ^4,5,9^. There may be similar toxins in the environment that mimic the effects and properties of biliatresone ^10^. In our institutional experience, we noticed significant annual fluctuation in both biliary atresia cases as well as severity of red tide outbreaks in the Southwest Gulf of Mexico (Figure 1) prompting the hypothesis that there could be a relationship between the two phenomena. There have been few studies exploring the association between environmental toxins and development of BA in humans ^10^. Red tide blooms, caused by the dinoflagellate *Karenia brevis*, occur in the late summer and early fall but can last days, weeks or months ^11^. *K. brevis* produce brevetoxins, which are lipophilic, cyclic polyethers ^12–16^ that cause severe morbidity and mortality among wildlife ^12,15,17–26^, developmental anomalies in fish embryos ^27,28^ and respiratory toxicity, gastrointestinal symptoms, and neurotoxicity in humans ^29–32^. Brevetoxins impair immune system function ^13^ and disturb DNA structure ^27,33^. Across multiple species, brevetoxins are concentrated in the muscles, intestine and notably, the liver, with primary elimination via biliary excretion ^13,15,16,19,21,23,33,34^ and with toxic effects that appear to persist beyond a single exposure ^13^.

**Figure 1:**
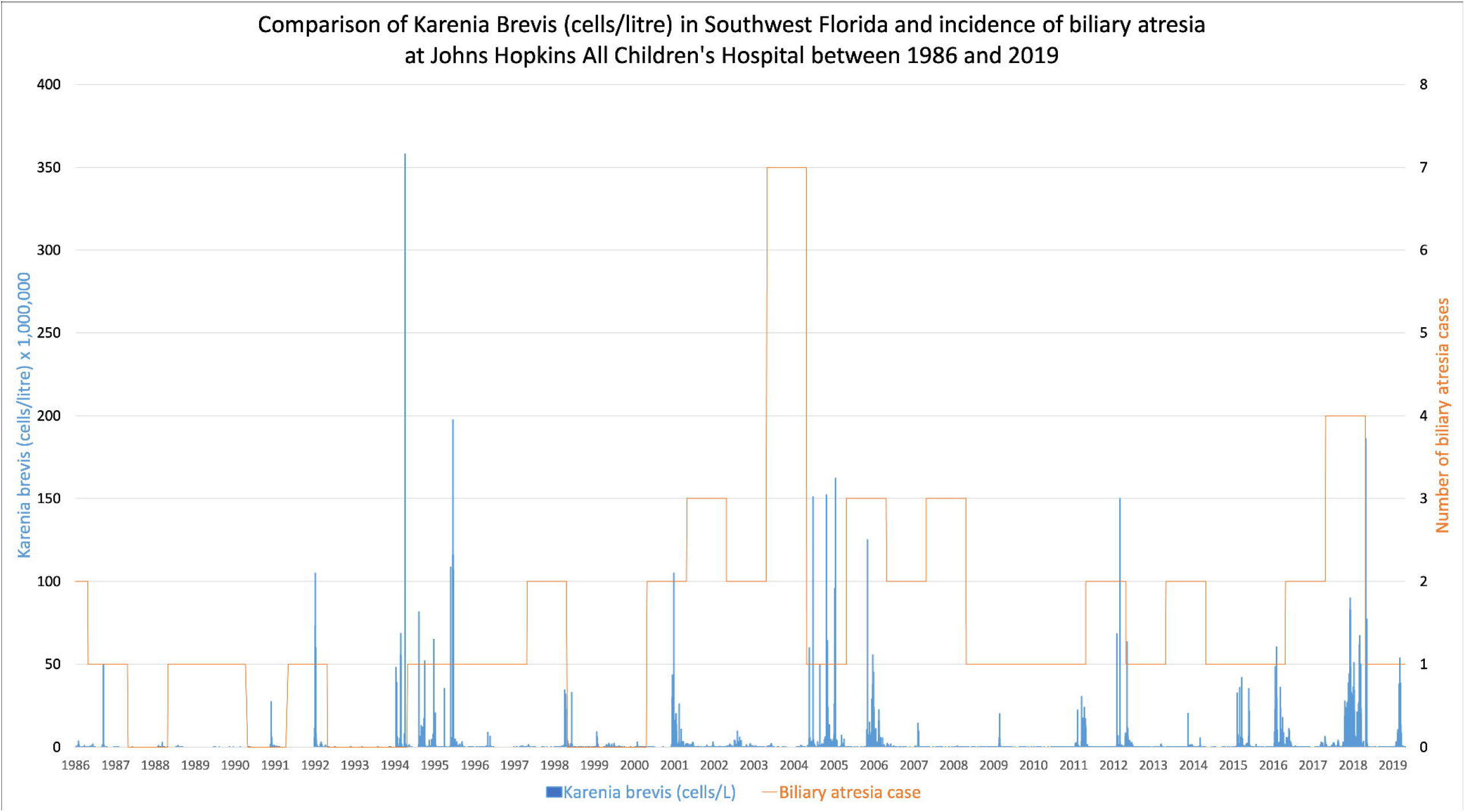
*K. brevis* (cells/liter) levels as collected and reported at multiple timepoints per annum by the Florida Fish and Wildlife Conservation Commission with superimposed incidence of biliary atresia cases at Johns Hopkins All Children’s Hospital between 1986 and 2019.

We therefore conducted a case-control study to evaluate whether maternal exposure to *K. brevis* (cells/liter) during gestation was associated with BA in offspring.

## METHODS

### Subjects and study design

This was a single-center, hospital-based, case-control study of patients seen at Johns Hopkins All Children’s Hospital from 10/2/2001 to 12/23/2019. The electronic medical record was queried for prespecified ICD-10 and ICD-9 codes indicative of BA: Q44.2, Q44.5, Q44.0, Q44.1, Q44.3, Q44.7, 751.61. Cases were infants with a diagnosis of BA confirmed after independent review of the medical records and liver biopsy pathology reports. For each case, 4 controls were individually matched by birth month and year. Controls were randomly selected from a pool of patients without a diagnosis of BA who were admitted to the hospital during the study period, and who were diagnosed with common pediatric respiratory infections (bacterial or viral), foreign body ingestions, injuries, gastrointestinal reflux, asthma or common dermatologic diseases of childhood without any documented chronic or genetic disorders or congenital anomalies.

Cases and controls were excluded if they were diagnosed with Alagille syndrome, congenital heart disease with cholestasis, Down syndrome with cholestasis, neonatal cholestasis, congenital cystic adenomatoid malformation, choledochal cyst, progressive familial intrahepatic cholestasis, congenital syphilis, VACTERL syndrome, gastroschisis with cholestasis, genetic abnormalities, cryptogenic cirrhosis, Kabuki syndrome, cystic fibrosis, Caroli syndrome, Noonan syndrome, Hirschsprung with cholestasis, and other ductal problems not deemed to be BA upon pathology review. Additionally, cases and controls were excluded if they were non-Florida residents. There were no biological relationships among cases or controls. The study was approved by Johns Hopkins All Children’s Hospital Institutional Review Board (IRB00252758) prior to initiation.

### *K. brevis* exposure assessment

Zip-code level *K. brevis* exposure was mapped to the patient-level for each case and control using water concentration levels routinely measured and reported in Gulf coast counties by the Florida Fish and Wildlife Conservation Commission (FWC). Zip-code level exposure data from the FWC were exported from the FWC database in 28-day increments based on a full term 40-week gestation extrapolated from the patient date of birth. Queries were exported as comma separated value spreadsheets and converted to shapefiles based on latitude and longitude using the ArcCatalog (10.7.1) “Add XY Data” tool. The shapefiles were then projected to NAD 1983 HARN Florida GDL Albers using ArcCatalog’s “Project” tool. Using patient zip codes as a proxy for residence, both 10-mile and 50-mile radii were drawn using the “Buffer” tool to assign an individual-level *K. brevis* exposure to each patient. Data were then exported for statistical analyses.

### Covariates

Gender and race/ethnicity, based on self-report, were collected at hospital admission. Patients were categorized according to whether they lived within an area designated as coastal by independent determination of whether the patient’s zip code directly abutted Tampa Bay or the Gulf of Mexico. To analyze seasonality of birth, dates of birth were specified as summer (June 21 to September 21), fall (September 22 to December 20), winter (December 21 to March 19) or spring (March 20 to June 20).

### Statistical analyses

Demographic characteristics were summarized with frequencies and percentages for categorical variables. Median gestational *K. brevis* exposure within a 10- and 50-mile radius of the household zip code was calculated and reported with interquartile ranges and minimum to maximum values. Maximum gestational toxin exposure was calculated and categorized using clinically relevant cut-points for descriptive purposes (i.e., none/not present: <1000 cells, low: >1,000-99,000, medium: ≥100k – 999k, high: ≥1 million), as routinely stratified by the FWC. Statistical comparisons between cases and controls were performed using Mann-Whitney *U* test for continuous variables and Chi-square or Fisher’s exact test for categorical variables as appropriate.

Conditional multivariable logistic regression models matched for age were constructed to evaluate the adjusted association between maximum *K. brevis* exposure per trimester at the 10- and 50-mile radius in mothers and odds of BA in neonates. The primary independent variable, maximum gestational *K. brevis* exposure, was dichotomized into none/not present/low exposure (<1000 cells/liter to 99,000 cells/liter) and medium/high exposure (≥100k cells/liter and above). To arrive at the final multivariable models, gender, race/ethnicity, coastal residence, and season of birth were tested as potential confounders of the relationship between *K. brevis* exposure and BA using the 10% criterion for confounding. Multi-categorical covariates, including race/ethnicity and season of birth, were dichotomized for statistical efficiency. Missing data were not imputed. Adjusted odds ratios and 95% confidence intervals were calculated. Two-sided *p*-values < .05 were considered statistically significant. All analyses were conducted with Stata/SE Version 17.1 (StataCorp LLC, College Station, TX.).

### Sample Size Considerations

Power was examined using PASS 16 Power Analysis and Sample Size Software (2018, NCSS, LLC; Kaysville, Utah, USA, ncss.com/software/pass). Given the lack of evidence on the relationship under study, a population standard deviation of 1.5 in the exposure as well as an R^2^ of 0.2 between exposure and covariates was assumed based on clinical expertise. With these considerations and a sample size of 38 cases matched to 152 controls in a 1:4 ratio, the study had an estimated 90% power to detect an odds ratio of 2.0 at a significance level of .05 utilizing conditional multivariable logistic regression.

## RESULTS

### Study population characteristics

Upon query of the EMR during the study period, 109 unique patients over 521 patient encounters were identified. After inclusion/exclusion criteria were applied, there were 38 cases of BA for which 152 controls were selected. Demographic characteristics of the study population are presented in Table 1. The distribution of gender differed between cases and controls; 55.3% (n=21) of cases were female and 64.5% (n=98) of controls were male (*p*=.03). Race/ethnicity, season of birth, and coastal residence did not vary significantly between cases and controls (*p*≥ .05 for all).

**Table 1.** Demographic characteristics of cases and controls

| **Table 1.** Demographic characteristics of cases and controls |  |  |  |
| --- | --- | --- | --- |
| Variable, Statistic | Cases (n = 38) | Controls (n = 152) | *p* value |
| **Gender, n(%)** |  |  |  |
| Male | 17 (44.7) | 98 (64.5) | .03 |
| Female | 21 (55.3) | 54 (35.5) |  |
| Unknown/Not Reported | 0 (0.0) | 0 (0.0) |  |
| **Race/Ethnicity, n(%)** |  |  |  |
| White | 24 (63.2) | 86 (56.6) | .3 |
| Black | 5 (13.2) | 37 (24.3) |  |
| Hispanic | 5 (13.2) | 17 (11.2) |  |
| Asian, Other | 3 (7.9) | 5 (3.3) |  |
| Unknown/Not Reported | 1 (2.6) | 7 (4.6) |  |
| **Season of birth, n(%)** |  |  |  |
| Summer | 7 (18.4) | 33 (21.7) | >.9 |
| Fall | 15 (39.5) | 51 (33.6) |  |
| Winter | 8 (21.1) | 35 (23.0) |  |
| Spring | 8 (21.1) | 33 (21.7) |  |
| Unknown/not reported | 0 (0.0) | 0 (0.0) |  |
| **Coastal Residence, n(%)** |  |  |  |
| Yes | 18 (47.4) | 89 (58.6) | .2 |
| No | 20 (52.6) | 63 (41.5) |  |
| Unknown/not reported | 0 (0.0) | 0 (0.0) |  |
| Percentages may not add up to 100% due to rounding |  |  |  |

### *K. brevis* exposure in cases and controls

The median gestational exposure at the 10-mile radius was 0 cells/liter among both controls (IQR: 0 to 1000.8 cells/liter) and cases (IQR: 0 to 0 cells/liter; *p*=.2). As seen in Figure 2, at the 50-mile radius, the median gestational exposure was 2000 cells/liter among both controls (IQR: 666.5 to 742500 cells/liter) and cases (IQR: 416.5 to 1063500 cells/liter; *p* >.9). Median gestational *K. brevis* exposure at the 50-mile radius peaked at 6 months of gestation (median: 4500 cells/liter, IQR: 333 to 2010000 cells/liter) among cases and 9 months (median: 6000 cells/liter, IQR: 667 to 386000 cells/liter) among controls.

**Figure 2:**
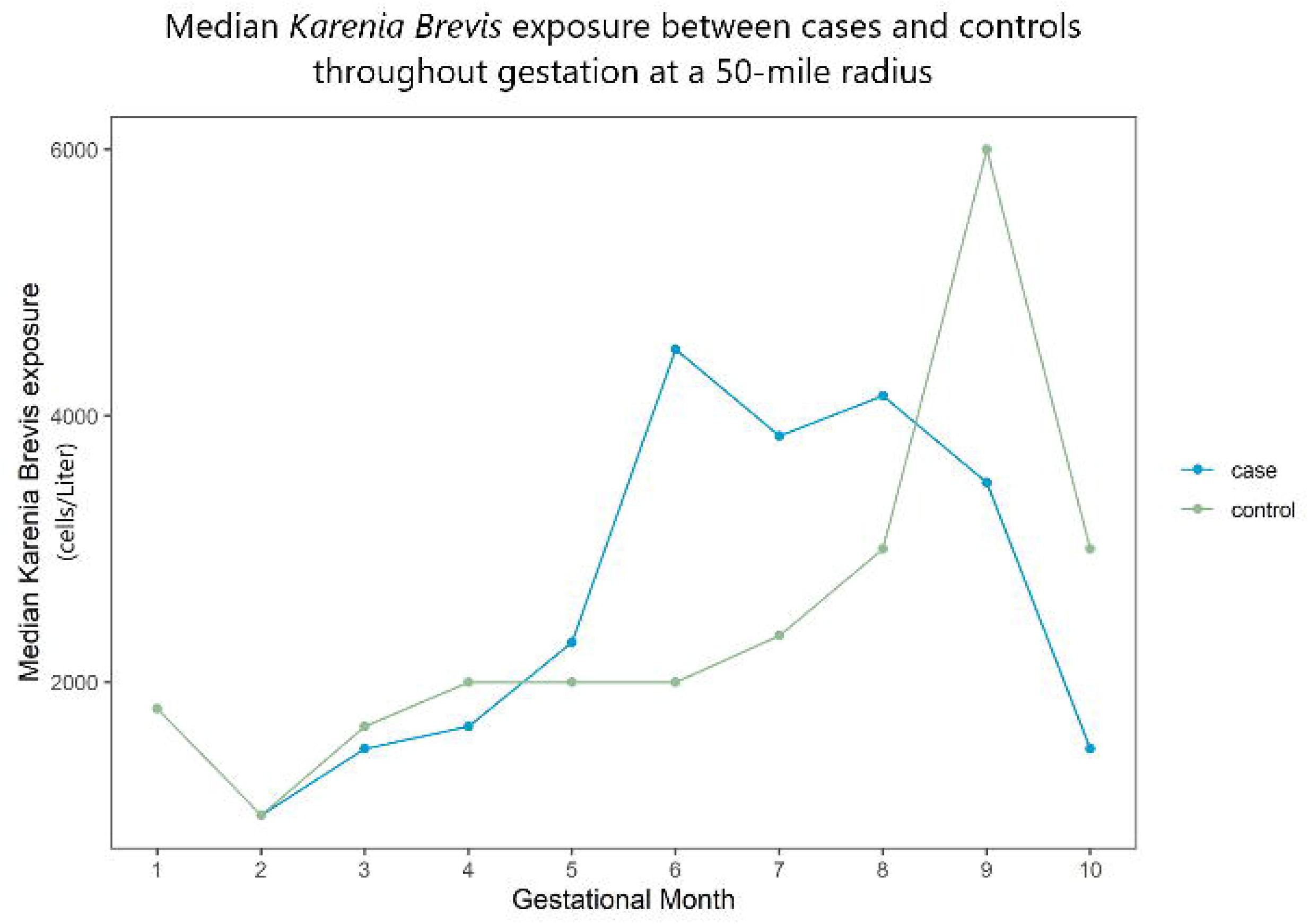
Median *K. brevis* (cells/liter) exposure for cases and controls throughout gestation at a 50-mile radius.

The distribution of maximum gestational *K. brevis* exposure at the 10-mile radius differed between cases and controls. Specifically, a greater proportion of controls had high *K. brevis* exposure (27.6%, n=42) in comparison to cases (18.4%, n=7) while a greater proportion of cases had no *K. brevis* exposure (63.2%, n=24) in comparison to controls (37.5%, n=57; *p*=.04). Maximum gestational *K. brevis* exposure at the 50-mile radius did not vary between cases and controls (*p*>.9). As seen in Figure 3a and 3b, when stratified by trimester, maximum *K. brevis* exposure at the 10- and 50-mile radius did not vary significantly between cases and controls (*p*≥ .05 for all).

**Figure 3:**
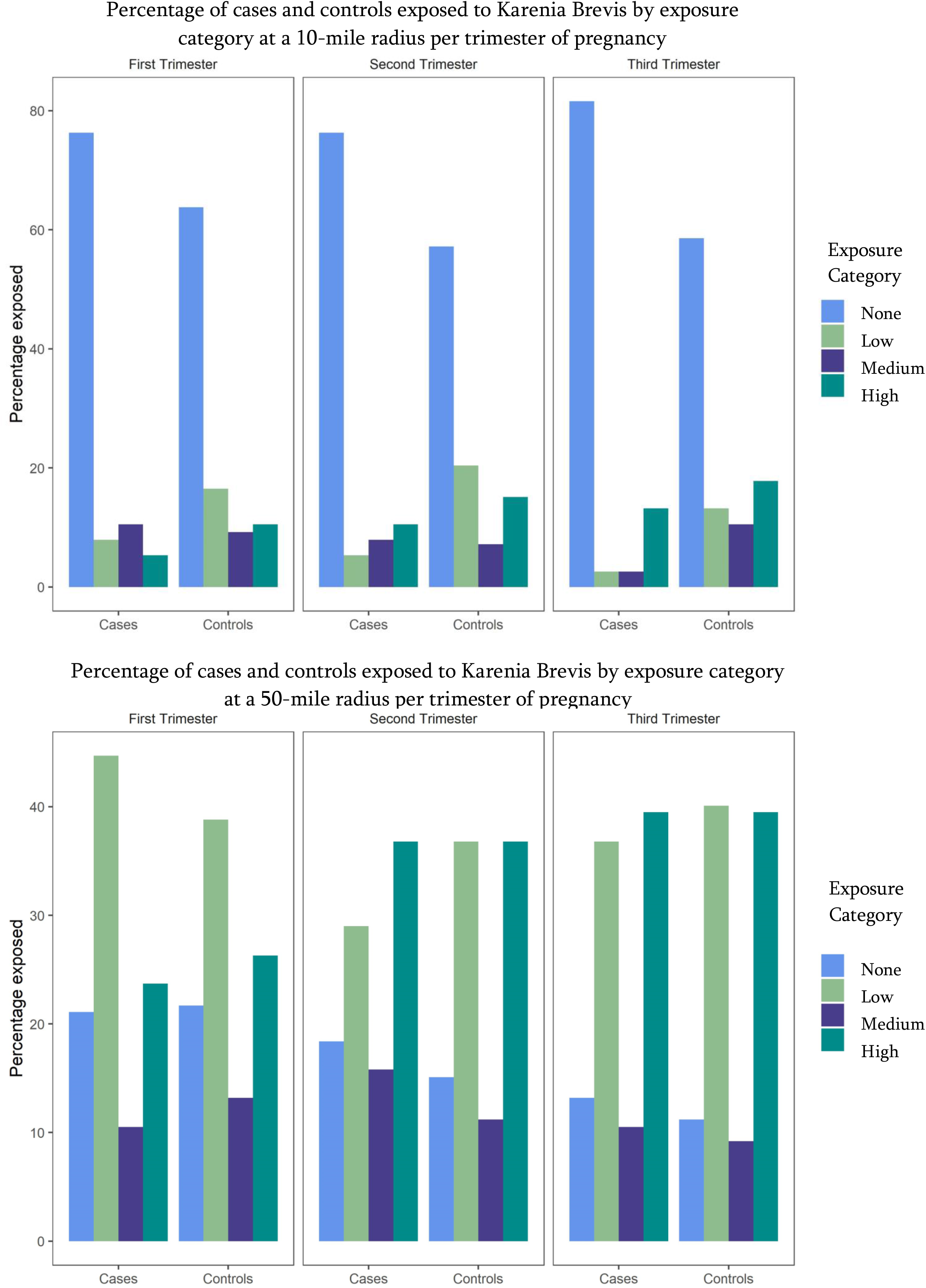
Percentage of cases and controls exposed to *K. brevis* as stratified by exposure category at a 10- and 50-mile radius. Exposure categories defined as none if <1,000 cells/L, Low if ≥ 1,000 to 99,999 cells/L, medium if ≥100,000 to 999,999 cells/L and high if ≥ 1,000,000 cells/L.

### Association between *K. brevis* and biliary atresia

Gender, race/ethnicity, coastal residence, and season of birth were observed as confounders of the relationship between *K. brev*is and BA according to the 10% criterion for confounding and were therefore included in the final multivariable models. As seen in Figure 4, there was not a statistically significant association between odds of BA and maximum *K. brevis* exposure per trimester at the 10- and 50-mile radius after adjustment for the aforementioned confounders. Notably, the direction of the adjusted odds ratio was less than one for all models except the second (OR: 3.2, CI: 0.5 to 20.4) and third trimester (OR:1.3, CI: 0.2 to 8.4) at the 50-mile radius.

**Figure 4:**
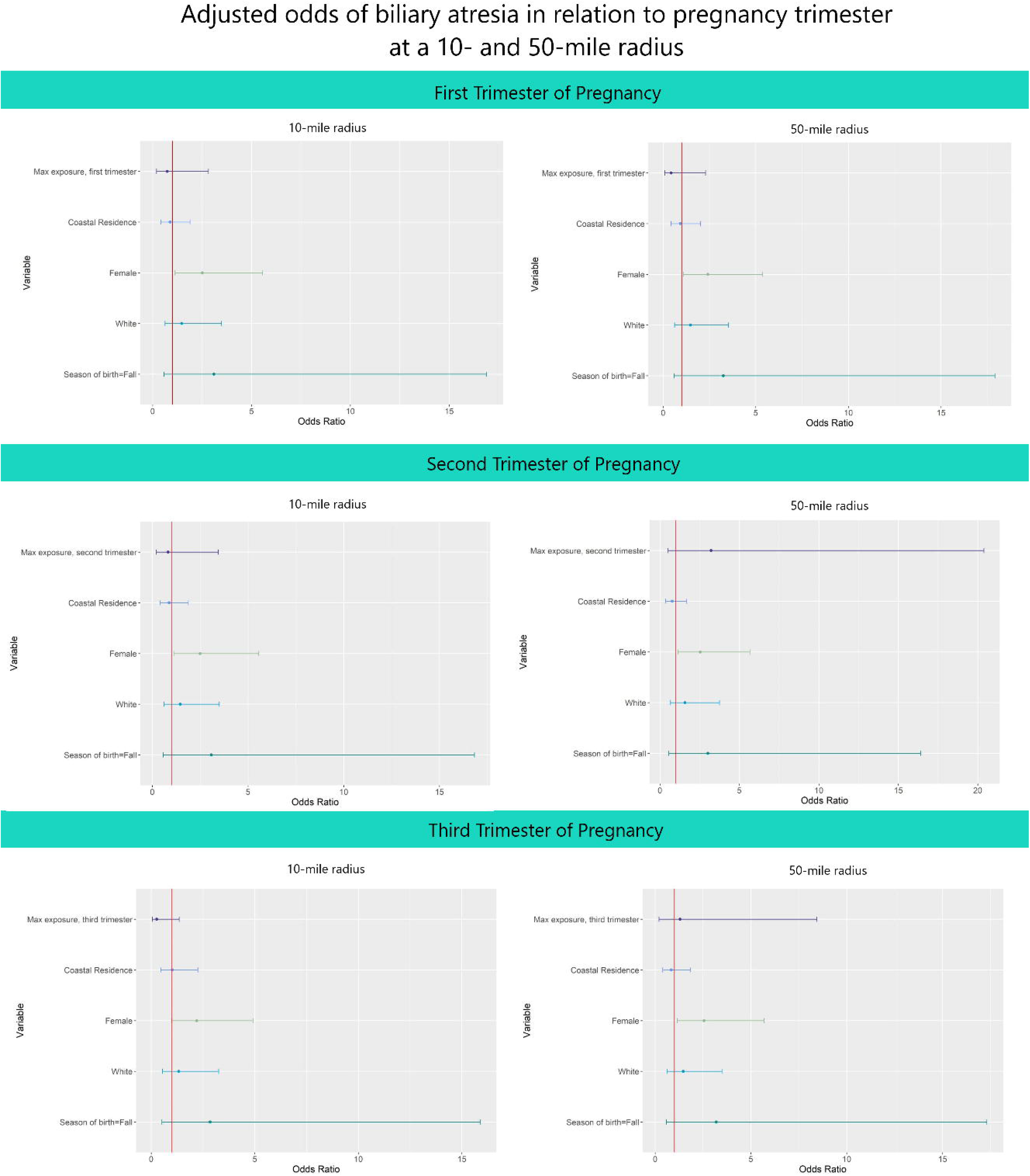
Association between biliary atresia and maximum *K. brevis* exposure per pregnancy trimester at a 10- and 50-mile radius.

## DISCUSSION

We evaluated whether an association between gestational *K. brevis* exposure and BA exists and observed no significant association using these data. The incidence of BA is increasing over time, with conflicting data regarding seasonality and risk factors ^35,36^. Our study did not account for the many proposed, yet not definitive, genetic, environmental or infectious confounders. Our cohort of patients did not demonstrate inter-group differences in birth seasonality, coastal residence or race/ethnicity; however, more cases were female.

There are a number of caveats to the results we obtained. We used *K. brevis* levels at 10- and 50-mile radii, extrapolated from the zip code of primary residence, as a surrogate for brevetoxin exposure. Monitoring *K. brevis* levels alone may not be the optimal predictor of red tide toxin (brevetoxin) exposure ^37^, although it is generally accepted that *K. brevis* levels of less than 1,000 cells/Liter are considered background levels while levels greater than 100,000 cells/Liter are associated with respiratory symptoms in humans and mortality among murine wildlife ^38^.

Brevetoxins have been detected in a variety of mammalian body fluids such as urine and serum using competitive enzyme-linked immuno-sorbent assay (competitive ELISA) ^39^. In humans, aerosolized brevetoxin has been quantified in scientific studies using an inhalable dust sampler and nasal swabs ^40^ though this is not routinely performed.

*K. brevis* levels can fluctuate daily depending on various meteorological conditions such as temperature, wind speed, sunlight, and rainfall ^41^. Brevetoxins can be aerosolized at the wind/water interface and transported up to 6.4 km inland ^32^ and changes based on its concentration in the water, wind speeds and wind direction ^42^. There exist a variety of possible direct exposures to brevetoxins including the consumption of contaminated shellfish, fish or seawater ^24^. Patient-level exposure to *K. brevis* was extrapolated from exposure at the community level in monthly intervals, but the true degree of daily exposure to brevetoxin, whether by direct ingestion or inhalation, was not measured in this study and is not routinely measured in humans. Patients from inland zip codes may travel to the beaches at various time points during red tide, making their true exposure difficult to quantify. Additionally, it appears that even respiratory inhalation can result in systemic distribution that can be long lived as evidenced by intratracheal administration of brevetoxin to mice which led to variable absorption, though about 20% of the initial dose was retained in the lung, liver and kidney beyond 7 days ^43^. Among turtles, brevetoxin can be stored and accumulated in multiple organs and passed on to offspring through the egg yolk, showing that brevetoxins can affect marine wildlife even after a red tide bloom has dissipated ^13^. It remains unclear whether brevetoxins behave in a similar way in humans but may be worth studying further given the detrimental effects in murine wildlife.

Overall, at a 10-mile radius from the zip code of residence, it appeared that our patient cohort had minimal exposure to *K. brevis* (cells/Liter) throughout gestation. Interestingly, at a 50-mile radius, the median gestational exposure for cases peaked at 6 months while for controls the peak occurred at 9 months. Recent studies explore the influence of early in utero events and the development of BA ^36^, and so it is worth noting that the timing of a toxic insult may be more important than absolute exposure.

In Southwest Florida, there is an annual fluctuation in the incidence of red tide blooms. Owing to the increasing severity of red tide over time, there is mounting concern for human contribution through water pollution carrying industrial effluents, organic pollutants, and excess nutrients on which *K. brevis* algae thrive ^25^. It is suggested that increasing water temperatures through climate change may exacerbate red tide ^44^. This study is unique in its targeted assessment of a specific, biologically plausible, environmental toxin and its association with BA, and though none was found using the study methodology, further studies quantifying brevetoxin levels in maternal and neonatal biological samples could be useful in determining association with disease. Continued inquiry into other environmental toxins and pollutants as a potential cause of BA is needed.

The lack of a significant association between K brevis exposure and development of BA must be carefully interpreted in the context of the limitations inherent to its design as a case control study. Controls were selected after an independent review of the electronic medical record and patients were excluded if they had any documented significant congenital anomalies, however, such diagnoses may exist outside of the documentation available within our hospital network. It is possible that certain common pediatric respiratory diagnoses may be aggravated by red tide toxin and may have been included in the control group; therefore, a large number of controls were selected to mitigate this potential risk. As a specialized children’s hospital, we serve a local area with multiple affiliates, however, patients with BA may have been diagnosed and treated elsewhere and thus, not captured in our study population. Additionally, as a single-institutional experience, our patient population may not be generalizable.

## CONCLUSIONS

In the population studied, there was no statistically significant association between gestational exposure of *K. brevis* (cells/Liter) at the 10- and 50-mile radius from the zip code of residence and increased odds of diagnosis with BA. Further studies into environmental factors or toxins are needed to elucidate the multifactorial pathophysiology of this complex disease.

## Data Availability

All data produced in the present study are available upon reasonable request to the authors

## ACKNOWLEDGEMENTS

We thank the Florida Fish and Wildlife Conservation Commission for graciously sharing their detailed records of red tide levels.

